# Disruption of the topological associated domain at Xp21.2 is related to gonadal dysgenesis: A general mechanism of pathogenesis

**DOI:** 10.1101/2020.03.25.20041418

**Authors:** Ana Paula dos Santos, Jakob A. Meinel, Cristiane dos Santos Cruz Piveta, Juliana Gabriel Ribeiro de Andrade, Helena Fabbri-Scallet, Vera Lúcia Gil-da-Silva-Lopes, Gil Guerra-Júnior, Axel Künstner, Frank J. Kaiser, Paul-Martin Holterhus, Olaf Hiort, Hauke Busch, Andréa Trevas Maciel-Guerra, Maricilda Palandi de Mello, Ralf Werner

## Abstract

Duplications of dosage sensitive sex-locus Xp21.2 including *NR0B1* have been linked to 46,XY gonadal dysgenesis (GD) and their effects are attributed merely to increase gene dosage of *NR0B1* (*DAX1*). Here we present a general mechanism how deletions, duplications, triplications or inversions with or without *NR0B1* at Xp21.2 can lead to partial or complete GD by disrupting the cognate topological associated domain (TAD) in the vincinity of *NR0B1*. Our model is supported by three unrelated patients: two showing a 287kb overlapping duplication at the Xp21.2 locus upstream of *NR0B1* containing *CXorf21* and *GK* and one patient having a large new triplication of Xp21.2 as the most likely cause of GD. Whole Genome sequencing uncovered the exact structural rearrangements of the duplications and the triplication. Comparison with a previously published deletion upstream of *NR0B1* revealed a common 35kb overlap between the deletion, our newly reported *NR0B1* upstream duplications and the triplication as well as all other copy number variations (CNVs) at Xp21.2 reported so far. This overlap contains a strong CCCTC-binding factor (CTCF) binding site representing one boundary of the *NR0B1* TAD. All three CNVs at Xp21.2 most likely disrupt this TAD boundary, which isolates *NR0B1* from *CXorf21* and *GK* and putatively results in *GK* and *CXorf21* enhancer adoption and ensuing ectopic *NR0B1* expression. As a result, the patients’ transcriptomes developed an intermediate expression pattern with both ovarian and testicular features and greatly reduced expression of spermatogenesis-related genes. This model not only allows better diagnosis of GD displaying CNVs at Xp21.2, but also gives deeper insight how spatiotemporal activation of developmental genes can be disrupted by reorganized TADs also in other rare diseases.

## Introduction

46,XY differences/disorders of sex development (DSD) with gonadal dysgenesis (GD) are characterized by a range of phenotypes from genital ambiguity to female genitalia caused by variable degrees of testicular failure. Genetic variants in more than 20 genes have been described as monogenic causes, several of these in a dose-dependent manner (Audi et al., 2018). Among such genes is *NR0B1* (*Nuclear Receptor Subfamily 0, Group B, Member 1*; also known as *DAX1*), located within a dosage-sensitive 160kb spanning sex reversal (DSS) region at Xp21.2 (Bardoni et al., 1994). Since its first description several 46,XY GD patients with duplications including the entire *NR0B1* have been reported (Figure S1, Supporting Information) (Barbaro et al., 2008; Barbaro, Cook, Lagerstedt-Robinson, & Wedell, 2012; Barbaro et al., 2007; Dong et al., 2016; Garcia-Acero et al., 2019; Ledig et al., 2010; White et al., 2011). In mice, *Nr0b1* expression starts in the genital ridge of both sexes at the same time point as the expression of *Sry* (*sex determining region Y*), but is downregulated in the developing testis and persists in the ovary (Swain, Zanaria, Hacker, Lovell-Badge, & Camerino, 1996). It has been shown that high exogenous *Nr0b1* expression in transgenic mice retards testis formation. Coexpression with a weak *Sry* allele even resulted in complete sex reversal (Swain, Narvaez, Burgoyne, Camerino, & Lovell-Badge, 1998) resembling the *NR0B1* duplications in humans that cause 46,XY GD (OMIM #300018). Thus, *NR0B1* is the most plausible candidate gene for 46,XY GD in the DSS region. In humans, *NR0B1* inactivating mutations cause X-linked congenital adrenal hypoplasia (AHC) with hypogonadotropic hypogonadism (OMIM #300200), but boys have intact testicular development at birth.

*Nr0b1* is known to directly repress *Sox9* (*Sry - box 9*) expression rather than expression of upstream targets, such as *Sf1* (*steroidogenic factor 1*; also known as *NR5A1*) or *Sry*, whose expression remains unchanged under NR0B1 overexpression (Ludbrook et al., 2012). *Sox9* is important for Sertoli cell differentiation and its activation is *Sf1* dependent (Figure S2, Supporting Information) (Ludbrook et al., 2012). NR0B1 exerts its anti-testis effect through disruption of both, SF1 DNA-binding and *Sox9* enhancer activation in a dose-dependent manner (Ludbrook et al., 2012). However, while high *Nr0B1* expression is incompatible with testicular development (Swain et al., 1998) it is later required for preserving spermatogenesis (Yu, Ito, Saunders, Camper, & Jameson, 1998) implying it itself must be precicely regulated in the testis.

Despite considerable advances in the understanding of sex development, the genetic etiology of many 46,XY GD patients still remain unclear (Audi et al., 2018; Bashamboo, Eozenou, Rojo, & McElreavey, 2017). Besides unidentified genes in the gonadal developmental pathway, this may be due to a neglect of non-coding and regulatory elements in the molecular analysis of these patients so far. Whole genome sequencing (WGS) transcends this paradigm by providing detailed data on the complete patient genome. Here we describe three 46,XY patients with GD have been raised as females and for whom we did not find any protein-coding variants in the genome considered as pathogenic for the phenotype. Still, a transcriptome analysis of gonadal tissue revealed expression of both testis and ovary related genes, such as *SOX9* and *FOXL2*, respectively, and showed a greatly reduced expression of spermatogenesis pathways (Figure S3, Supporting Information). Remarkably, all three patients carry Xp21.2 copy number variations (CNVs) with a 287kb overlapping region upstream of *NR0B1*. Comparison with a published deletion at Xp21.2 (Smyk et al., 2007) narrowed this region to 35kb. Based on publicly available chromatin immunoprecipitation (ChIP) and chromatin conformation capture (3C, Hi-C) data (Y. Wang et al., 2018) we propose that this common region includes a boundary element of a topologically associating domain (TAD). The genomic duplications or the triplication and the deletion upstream of *NR0B1* disrupts this TAD and the association of the genes and their specific regulatory elements therein. This would result in upregulation of *NR0B1* expression which causes suppression of *SOX9*, disrupting Sertoli cell differentiation and successful testis-development (Chaboissier et al., 2004).

## Patients and Methods

### Editorial Policies and Ethical Considerations

This study was approved by local Ethics Committees of the Universities of Campinas (*CAAE 24972513*.*5*.*0000*.*5404*) and Lübeck (*AZ 17-219*). Written informed consent was obtained from the patients and/or their parents/legal guardians.

### Patients

Three unrelated patients from Brazil and Germany were investigated in the study.

### Patient 1 (P1)

The patient was referred to the DSD clinic, State University of Campinas at age of 5 days, due to genital ambiguity. She was born at term after an uneventful pregnancy with birth weight of 3,410 g and a length of 49 cm. She had a 0.5-cm phallus, a single perineal opening, partially fused labioscrotal folds and non-palpable gonads (External Masculinization Score - EMS = 4) (Ahmed, Khwaja, & Hughes, 2000). Abdominal ultrasound revealed no abnormalities and genitography showed a urogenital sinus. The karyotype was 46,XY and fluorescence in situ hybridization (FISH) displayed no 45,X cell line. Laparoscopy showed absence of uterus, likewise the left gonadal tissue was absent and there was a right dysgenetic testis with some areas of fibrous tissue surrounding immature seminiferous tubules without spermatogonia. When she was 1 month old, hormonal evaluation revealed high FSH (24 IU/l; normal range: 1.5 to 12.4 IU/l) and LH (10 IU/l; NR: 1.7 to 8.6 IU/l) levels and low testosterone (0.2 ng/ml; NR: 2.86 to 8.10 ng/ml), suggestive of testicular failure due to partial GD.

### Patient 2 (P2)

First presentation in the University DSD center in Kiel and Lübeck occurred at the age of 17 years and 2 months due to primary amenorrhea and pubertal delay. The girl previously had a dysgerminoma which was removed 3 months earlier. Tanner stages were B4, P4-5. However, breast development started only at the age of 16 years, a few months before clinical diagnosis of the dysgerminoma. External genitalia were completely female without clitoromegaly and a uterus with tubular configuration was present. The karyotype was 46,XY. Hormonal evaluation revealed basal LH 53 IU/l and FSH 94.3 IU/l increasing to 200 IU/l and 128 IU/l, respectively, 30 min following 60 µg/m^2^ GnRH i.v.. Plasma estradiol was 9 pg/ml (prepubertal for girls) and testosterone 123 ng/dl (mid-pubertal for males) (both determined by LC-MS/MS) (Kulle, Riepe, Melchior, Hiort, & Holterhus, 2010). Plasma AMH was 0.71 µg/l and therefore extremely low. Therefore, the clinical diagnosis of GD was established.

#### Patient 3 (P3)

This patient was born at term after an uneventful pregnancy and genital status was described as unequivocal female. The family history is unremarkable, with three healthy siblings. Because of muscular hypotonia, a chromosomal analysis was initiated and revealed a 46,XY karyotype. On ultrasound, a small prepubertal uterus was seen, but the gonads were not visualized. Hormone analysis showed a prepubertal status with inhibin B below the threshold and anti-Mullerian hormone at 0.42 µg/l, which was considered low for age, compatible with a clinical diagnosis of gonadal dysgenesis. A laparoscopy was performed and the gonadal tissue removed. The histology revealed a gonadoblastoma of the right gonad, while the left side was purely stromal tissue.

### Multiplex ligation dependent probe amplification (MLPA)

To identify DNA gains and losses, multiplex-ligation dependent probe amplification (MLPA) assays were performed for P1 using two different kits: SALSA MLPA kit P185-C1 Intersex probemix and SALSA MLPA P334-A2 Gonadal Development Disorder probemix (MRC-Holland, Amsterdam, The Netherlands). The MLPA assay was carried out according to the standard protocol supplied by MRC-Holland. Data were analysed by comparison of the peaks obtained from the tested samples and those from controls using the software *Coffalyser*.*NET* (MRC-Holland, Amsterdam, The Netherlands).

### Chromosomal microarray analysis (CMA)

Chromosomal microarray analyses were performed using two different platforms. DNA samples from P1 and her mother were hybridised onto the GeneChip CytoScan® 750K Array Kit (Affymetrix Inc., Santa Clara, California, USA) and processed as recommended by the manufacturer. Array data were analysed using Chromosome Analysis Suite (ChAS) (Affymetrix®). Standard analysis adopted the following parameters: markers >25 for deletion; >50 for duplication, without filter size for copy number variations (CNVs). DNA of P2 and P3 was hybridized to an Agilent 180K comparative genomic hybridization array (aCGH) (Agilent Technologies, Inc) and compared to a pooled sample of 10 normal males. Data were analysed using *CytoGenomics* Software version 4.0.2.21 (Agilent). CNVs of both patients were analysed using the Database of Genomic Variants (DGV, Version CNV_DGV_hg19_v4, Toronto, Canada).

### Whole Genome sequencing (WGS)

WGS was performed to identify deleterious point mutations and indels as well as structural variations and to fine map the breakpoints of the duplications and triplication at Xp21.2 observed in the three patients. Sequencing libraries were constructed from 1.0 µg DNA per sample using the Truseq Nano DNA HT Sample preparation Kit (Illumina, USA) following the recommendations of the manufacturer. Genomic DNA was randomly fragmented to a size of approximately 350 bp by Covaris cracker (Covaris, USA). DNA fragments were blunted, A-tailed and ligated with the full-length adapter for Illumina sequencing with further PCR amplification. Libraries were purified using AMPure XP (Beckman Coulter, USA), analysed for size distribution on an Agilent 2100 Bioanalyser (Agilent Technologies) and quantified by q-PCR. Paired end sequencing of libraries was performed on Illumina HiSeq platforms (Illumina). Per sample more than 90 GB of raw data were obtained, resulting in an average genomic read depth of 30x.

### Bioinformatics

Sequencing reads were mapped to human reference genome version GRCh37/hg19 using Burrows–Wheeler Aligner (BWA)(Li & Durbin, 2009). Resulting mapping files were screened for duplicated reads applying Picard tools *MarkDuplicates* version 1.111 (Picard: http://sourceforge.net/projects/picard/). Split-reads and discordant paired-end alignments were extracted using *SAMtools* version 0.1.18 (Li & Durbin, 2009). SNPs and InDels were called using *HaplotypeCaller* as implemented in Genome Analysis Toolkit (GATK) version 3.8.0 (DePristo et al., 2011) with standard parameters. Detection of structural variations (SV) was performed using DELLY (Rausch et al., 2012). Variations were annotated using ANNOVAR (K. Wang, Li, & Hakonarson, 2010). All chromosomal positions in the paper are according to GRCh37/hg19.

### Breakpoint Sequencing

Primers at either end of the constructed breakpoint sequences were designed using Lasergene PrimerSelect (DNASTAR, Wisconsin, USA). Optimal annealing temperature was determined through gradient PCR and electrophoresis. Consequently, PCR setup was 35 cycles with 30 sec. at 95°C for denaturation, 30 sec. at primer specific temperature (Table S2, Supporting Information) for annealing and 1 min. extension at 72°C. Sanger Sequencing of amplicons was performed on a 3130 Genetic Analyzer (Applied Biosystems, Foster City, USA).

### Transcriptome Analysis

Transcriptome analysis was performed from 10 slices (10 µm) of formalin fixed paraffin embedded (FFPE) samples of the gonads of patients 1 and 2. Library preparation was performed employing the TruSeq stranded Total RNA kit (Illumina Inc.), and sequenced on an Illumina NovaSeq platform (150-nucleotide paired-end reads), obtaining 98 and 63 million reads for patient 1 and 2, respectively. Fastq reads were pseudo-aligned to the Ensembl cDNA sequence Grch38 transcriptome assembly in version 96 using kallisto (Bray, Pimentel, Melsted, & Pachter, 2016). Gene transcript read counts were aggregated to Ensembl Gene IDs for further analysis (Soneson, Love, & Robinson, 2015) and genes with an expression less than one transcript per million (TPM) in either sample discarded, resulting in 13005 expressed genes.

Patient transcriptomes from expressed genes were compared to testis and ovary tissue samples from the GTEx Database (Carithers et al., 2015) and the data set E-MTAB-6814 from ArrayExpress (Athar et al., 2019), a human RNAseq time-series of the development of seven major organs.

To compare transcriptome similarity, we performed a dimensionality reduction using t-distributed stochastic neighbor embedding (t-SNE) (van der Maaten, 2014) as implemented in the R library Rtsne (version 0.15) (Krijthe, 2015). The analysis is based on 641 samples and 5000 randomly selected genes, expressed in both patient tissues.

Differential pathway activity between samples was determined by a Gene Set Variation Analysis (GSVA) (Hanzelmann, Castelo, & Guinney, 2013), which calculates the relative enrichment of gene sets across individual samples using a non-parametric approach. For pathway and upstream transcription factor activity we used the hallmark gene set collection (Liberzon et al., 2015) and the TF-target relationship from Schacht et al (2014) (Schacht, Oswald, Eils, Eichmuller, & Konig, 2014). The enrichment scores were tested for significant differences using a moderated t-test per time point (Ritchie et al., 2015).

## Results

### Patient 1

CMA revealed a ≈277kb duplication at Xp21.2 [30,580,693-30,857,187] that was supported by MLPA (Figure S4, Supporting Information). Analysis of the mother showed the same duplication. WGS confirmed this structural variation (SV) in the patient showing a 297kb duplication of *GK* (*glycerol kinase*), *CXorf21* (*chromosome X open reading frame 21*) and part of *TAB3* (*TGF beta activated kinase 1 binding protein 7, MAP3K7*). Duplication borders were apparent through increased read-depth and split reads mapping 297kb apart. Paired split reads were extracted and aligned to generate a continuous sequence of the breakpoints showing a tandem duplication, where intron 9 of *TAB3* is merged to the downstream region of *CXorf21* by a 27bp insert (Figure 1). The sequences at the breakpoint were confirmed by PCR and Sanger sequencing. Only two heterozygous missense SNVs were found in protein coding regions of genes related to 46,XY DSD (Table S1, Supporting Information). However, heterozygous pathogenic variants in both genes (*STAR*/*POR*) were excluded as putative cause of sex steroid biosynthesis deficiency.

**Figure 1.**
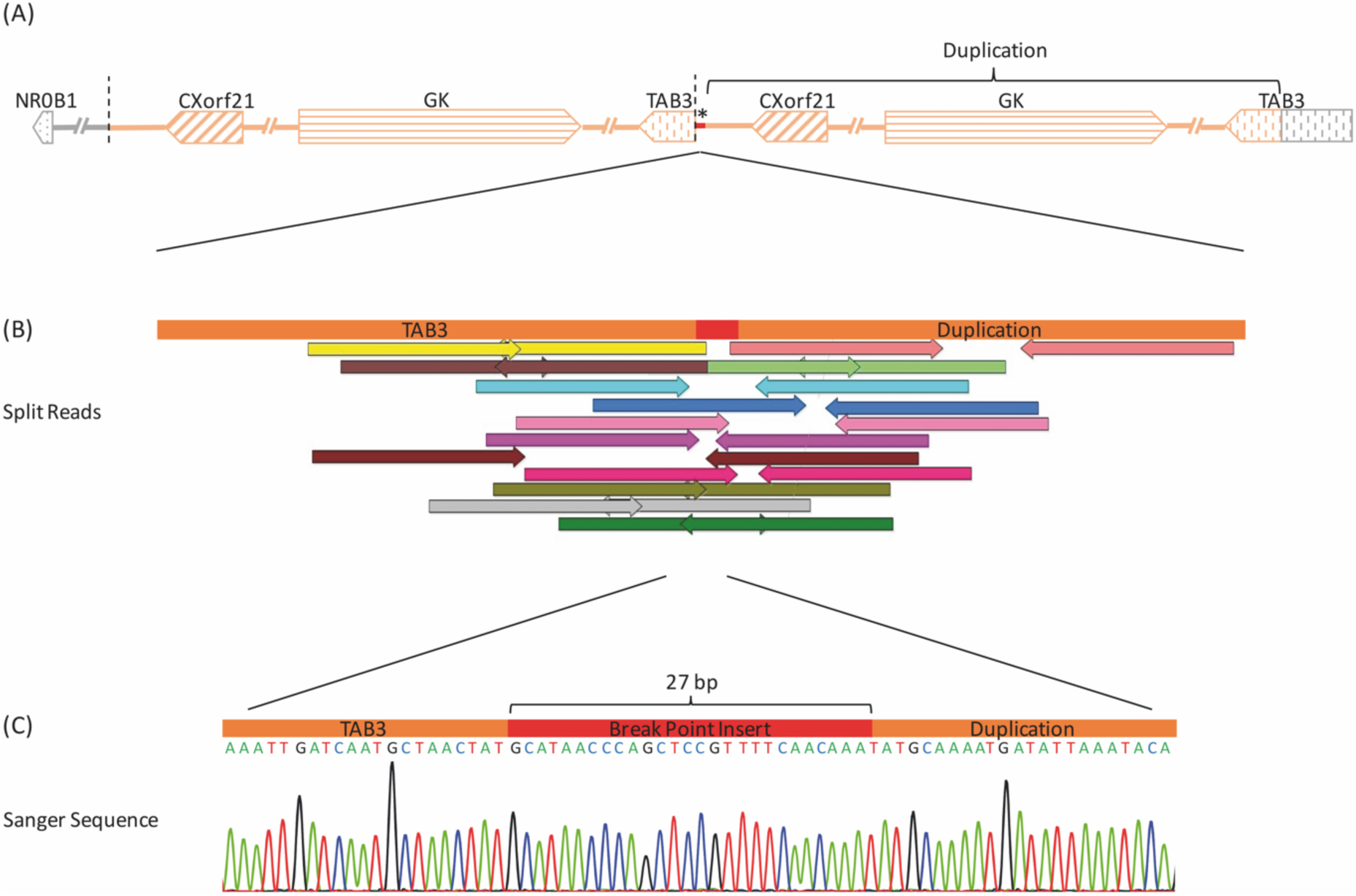
Structural Variation of Patient 1. (A) Overview of the copy number variation (CNV) in P1 upstream of the *NR0B1 gene* and its orientation within the Xp21.2 region. Arrows depict genes and their respective direction of transcription. The chromosome segment is drawn with the distal chromosome arm to the left and centromere to the right. The distances between genes are not to scale. The region chrX: 30,561,644-30,859,140 has been duplicated in a tandem manner, as indicated by the segments shaded orange. * marks a 27bp insert at the breakepoint. (B) Position of extracted and aligned split reads from genome sequencing crossing the breakpoint (chrX:30,859,140). Correspondingly coloured arrows of opposite direction belong to the same read pair. Read pairs have been mapped 297.5kb away from each other. (C) Verification of the breakpoint sequence by PCR and Sanger sequencing. Electropherogram shows a contiguous sequence from the *TAB3* gene to the breakpoint, an inserted sequence of 27bp (red) and the downstream sequence of the *CXorf21* gene at the beginning of the duplication.

### Patient 2

WGS revealed two major duplications and two small deletions at Xp21.2; one duplication of 389kb maps to a region downstream of *NR0B1* containing the *MAGEB* (*MAGE family member B*) genes 1-4 and a part of *IL1RAPL1* (*interleukin 1 receptor accessory protein like 1*). The second duplication (447kb), containing *CXorf21* and *GK* as well as the 3’-part of *TAB3*, maps upstream of *NR0B1*. Between the two duplications two small deletions of 2.7kb (Deletion I) and 2.2kb (Deletion II) flank an inverted region of 1.2kb (Figure 2; Figure S5, Supporting Information). As in P1, split read pairs at the borders of the four copy number variations (CNVs) were utilized to construct continuous breakpoint sequences and thus determine orientation and location of the inserted duplications. Both duplications are inserted upstream of *NR0B1* as described in Figure 2. Notably, the 447kb duplication encompassing *CXorf21, GK* and the *TAB3* fragment is inserted proximal to *NR0B1* in an inverted position. The 389kb duplication of *MAGEB1-4* and the *IL1RAPL1* fragment was inserted further upstream in the same orientation as the reference sequence. Breakpoint sequences were confirmed by Sanger sequencing (Figure 2) and copy number gains and losses predicted from WGS, were confirmed by aCGH (Figure S5, Supporting Information). WGS of the parents established that the SV was inherited maternally. Furthermore, only one rare variant (MAF<0.01) of unknown significance was found in the known DSD candidate gene *ZFPM2 (zinc finger protein, FOG family member 2)*. Although *ZFPM2* variants are associated with abnormalities in testis determination, the average population frequency of this SNV (dbSNP:rs202217256) is 0.004 across all populations and 0.006 in the Non-Finish European population according to the ExAc Browser (Lek et al., 2016), which indicates a rare polymorphism (Table S1, Supporting Information), rather than a relevant pathogenic variant leading to GD.

**Figure 2.**
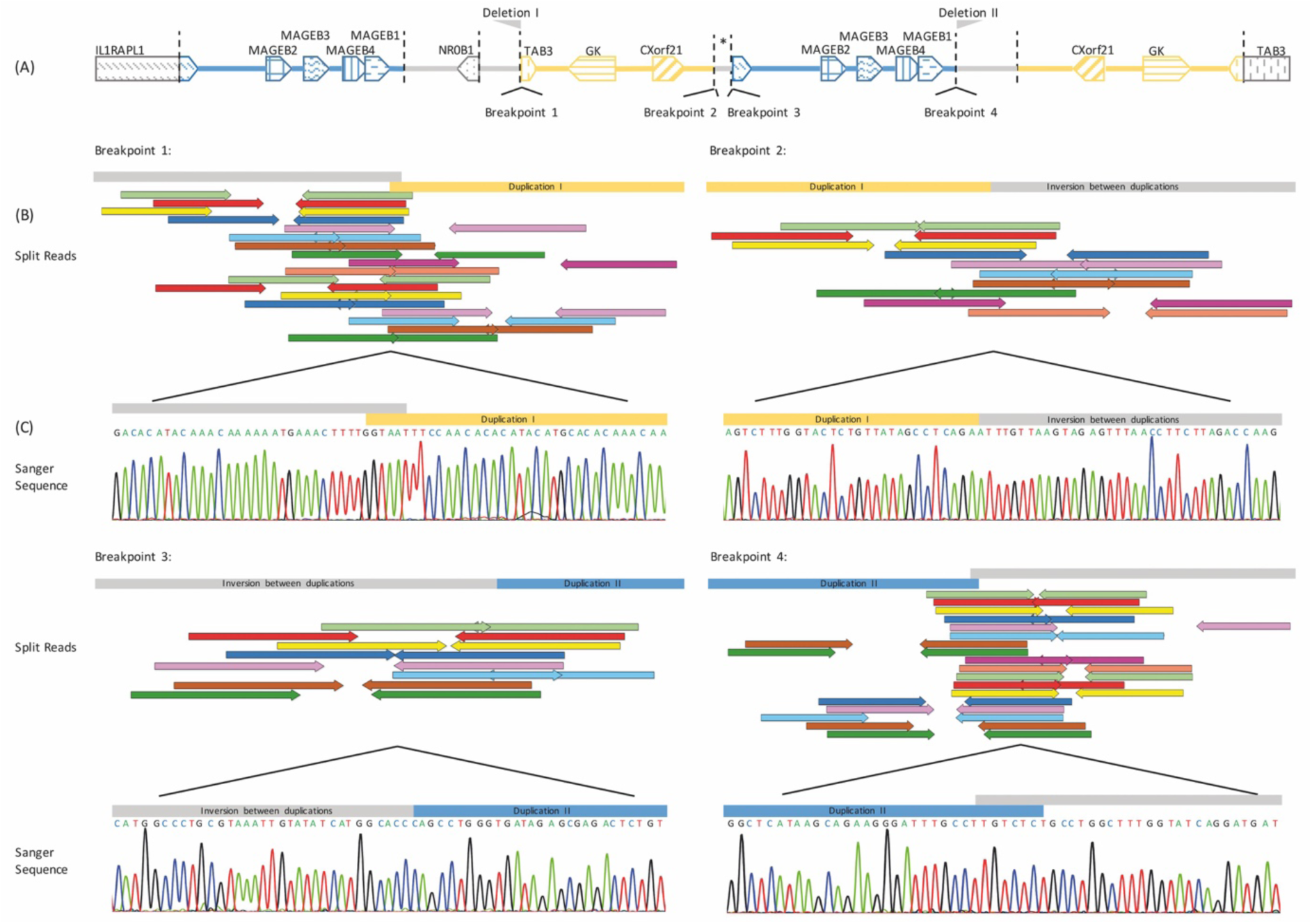
Structural Variation of Patient 2. (A) Overview of the structural variation upstream of the *NR0B1* gene. The genomic distances are not to scale. A 447kb duplication of *CXorf21, GK* and part of *TAB3* (yellow) originally mapping to 30,401,819-30,848,988 is inserted in an inverted manner between breakpoint 1 (bp1; chrX:30,336,745) and bp2 (chrX: 30,340,605) 9.25kb upstream of the *NR0B1* reading frame. * marks a 1.2kb piece of reference sequence inverted between bp2 and bp3 separating the two large duplications. Both duplications are flanked by deletions indicated by the grey flags denoted *Deletion I & II*. A 389kb duplication of the *MAGEB* genes 1-4 and part of *IL2RAPL2* (blue) originally mapping to chrX: 29,924,420-30,313,761 is inserted between breakpoint 3 (chrX: 30,339,452) and 4 (chrX: 30,342,785) in the same orientation as its reference sequence. (B) Shows position of extracted and aligned WGS split reads crossing the four breakpoints. Split reads mapped at varying distances apart for each breakpoint. Each pair was separated by at least >29kb. Sequences at either side of bp1 and bp4 showed homology, thus no exact definition of the breakpoint position was possible, as depicted by the overlap of the grey and yellow or blue bars. (C) Shows verification of sequences deduced from WGS reads through Sanger sequencing.

### Patient 3

P3 harbors a Xp21.2 triplication initially identified through qPCR copy number detection (Meinel, Dwivedi, Holterhus, Hiort, & Werner, 2019). In contrast to P1 and P2 this patients CNV also includes the *NR0B1* gene. An aCGH (Figure S6, Supporting Information) characterized the size of the triplication to be ≈1,22Mb, which was later rendered more precisely by WGS to be 1,24 Mb. The triplication includes the genes *MAGEB1-4, NR0B1, CXorf21, GK, TAB3* and part of *IL1RAPL1*. The triplicated segments are arranged in tandem to their template and are separated by a 49bp insert (ChrX:31,258,118-31,258,166) (Figure S7, Supporting Information). The insert originates form the *DMD* gene, which is located to the centromeric side of the CNV. Analysis of exome data of known DSD candidate genes revealed heterozygous missense SNVs, which all showed interpretations as benign or likely benign in ClinVar, except for one in *Estrogen Receptor 2* (*ESR2)* (Tabele S1, Supporting Information). The clinical significance of this SNV (dbSNP: rs367855747) has not been reported so far, however recently homo- and heterozygous *ESR2* mutations have been described in the context of 46,XY partial and complete GD (Baetens et al., 2018). The effect of this particular heterozygous SNV remains unclear, but in the context of widely agreed association of *NR0B1* copy number gains and 46,XY DSD this remains a subordinate factor in the etiology, if at all.

### Analysis of Topologically Associating Domains

SVs affecting boundaries of topologically associating domains (TADs) have been shown to alter the architecture and enhancer-promoter interactions within TADs with effects on development (Ibn-Salem et al., 2014; Lettice et al., 2011; Lupianez et al., 2015) and oncogenesis (Hnisz et al., 2016; Weischenfeldt et al., 2017). We analyzed the TADs in the *NR0B1* region identified by capture Hi-C from various human cell lines provided by the Penn State 3D genome browser (Y. Wang et al., 2018). Most cell lines show a clear compartmentalization of *NR0B1* into a TAD (referred to hereafter as *NR0B1* TAD) that is separated from the enhancer elements of the *CXorf21* and *GK* genes (Figure S8, Supporting Information).

When comparing the two duplications and the triplication observed in our patients with a previously published 257kb deletion in the *NR0B1* upstream region (Smyk et al., 2007), also associated with 46,XY GD, there is a 35kb overlap region among all four (Figure S1, Supporting Information). This region contains a strong binding site for CTCF (CCCTC binding factor), RAD21 and SMC3 (structural maintenance of chromosomes 3) (Figure S9, Supporting Information), which are involved in chromatin extrusion complexes and contact domain formation and are characteristic of TAD boundaries (de Wit et al., 2015; Sanborn et al., 2015). Thus, the binding sites in intron 2 of *CXorf21* most probably correspond to the *NR0B1* TAD boundary site that is observed in most cell types.

### Transcriptome Analysis of FFPE Gonadal Tissue

We compared the gonad transcriptomes from patient 1 and 2 to testis (n=156) and ovary tissue (n=133) transcriptomes that were obtained from the GTEx project and a human RNA-seq time-series from ArrayExresss containing tissue transcriptomes from seven major organs (brain (n=66), cerebellum (n=59), heart (n=46), kidney (n=37), liver (n=50), fetal ovary (n=18), fetal to infant testis (n=29) and teenager to old testis (n=10)) across different ages from fetal, infant, and teenager to adult and senior age. Lowly-expressed genes (TPM< 1) in the patients’ samples were discarded, leaving 13005 common genes. A t-SNE analysis reveals a strong tissue-specific clustering of all transcriptomes, independent of their experimental origin (Figure 3). Interestingly, fetal and young ovary and testis samples (pink and purple dots) cluster together, while adult ovary and testis samples (red and yellow/orange) significantly differ in their gene expression and cluster distinctly from any other organ. The samples of patient 1 and 2 are situated between the adult ovary and testis samples, yet close to patient samples that were reported by the GTEx project as having low spermatogenesis.

**Figure 3:**
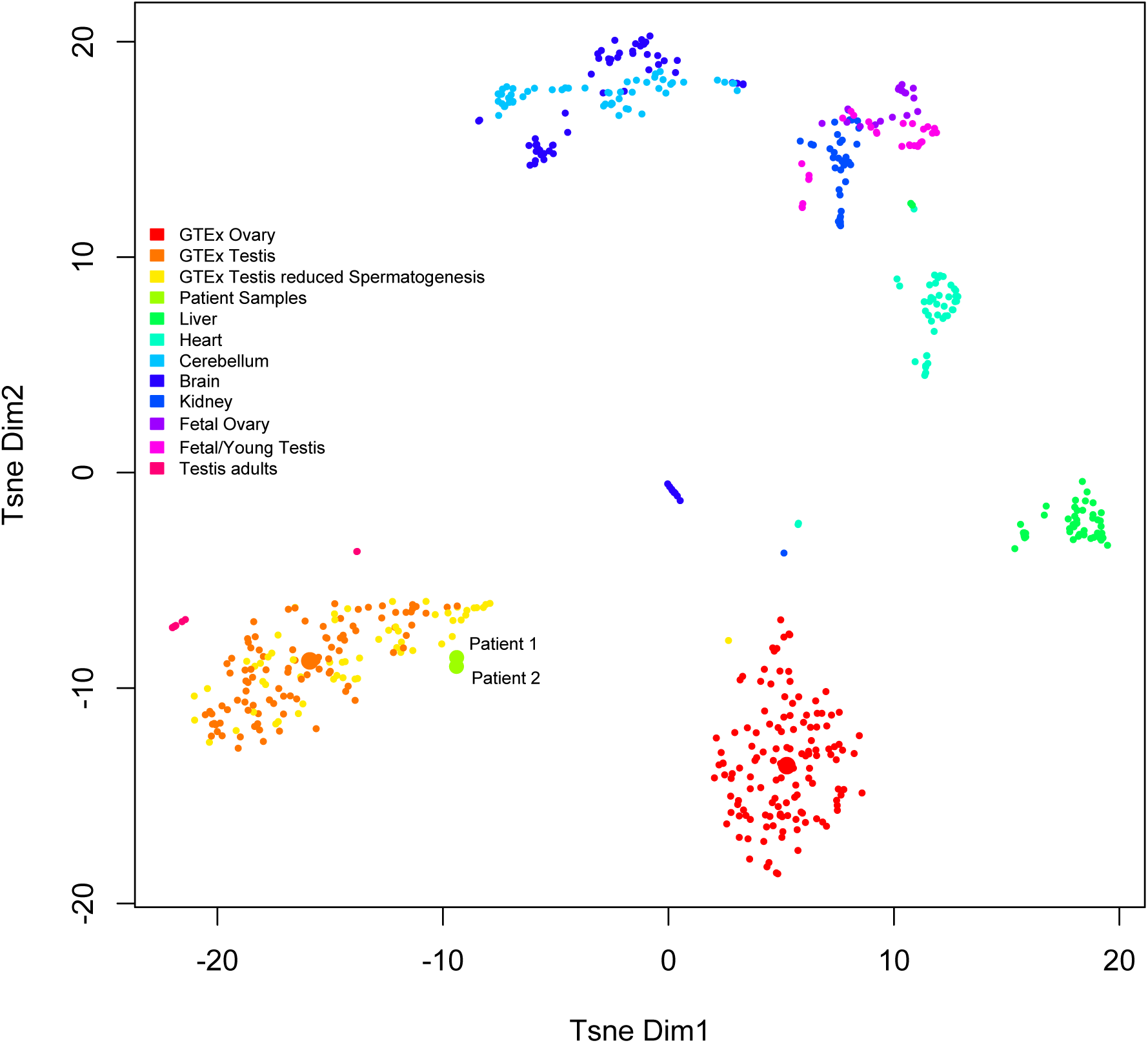
Comparison of patient gonad transcriptomes to healthy tissues. The two-dimensional t-SNE plot compares the transcriptomes of testis and ovary tissue from young and old healthy donors as well as from liver, heart, cerebellum, kidney and brain. The samples come from the GTEx database (red, orange, yellow points) as well as data set E-MTAB-6814 from ArrayExpress. t-SNE was calculated for 608 samples using randomly 5000 out of 13,005 genes expressed in either patient sample. Testis samples are marked as either orange or yellow, depending on whether they are reported to have normal or reduced spermatogenesis. The large orange and red sports among the testis and ovary tissue group mark the t-SNE the median expression of all tissue samples.

Figure S3 (Supporting Information) depicts boxplots of several sex determination genes from the GTEx ovary and testis samples used in Figure 3 and the two patients. Obviously, all genes are differentially expressed in the two tissues, but also between the patients and the ovary as well as the testis samples. For example, the *forkhead transcription factor FOXL2*, which is essential for ovary differentiation and maintenance, repressing the genetic program for somatic testis determination (Boulanger et al., 2014; Uhlenhaut et al., 2009), is highly specific to ovarian tissue, yet it is expressed at intermediate strength in the two patient samples with the expression in P2 exceeding that of P1. In contrast, the *Doublesex and Mab-3 Related Transcription Factor 1 DMRT1* is a transcription factor playing a key role in male sex determination and differentiation by controlling testis development and Sertoli cell maintenance (Ledig, Hiort, Wunsch, & Wieacker, 2012; Matson et al., 2011) is expressed in testis, but not in ovarian tissue. Yet, the patients’ gonads express this gene at an intermediate level.

This intermediate gene expression state is reflected by Gene Set Variation Analysis using the hallmark gene sets from the MSigDB (Liberzon et al., 2015) on 6 randomly selected testis and ovary tissues as well as the median tissue expression values from GTEx and the patient samples. The analysis shows how the latter have acquired expression programs from both tissues and cluster in between the ovary and testis samples. The most active pathway in testis is spermatogenesis. This pathway is not expressed in the ovary samples, yet low, but significantly higher in the patients. Interestingly, MYC signaling is present is both sample groups, but completely absent in the two patient samples (Figure S10A, Supporting Information). The latter finding is supported by comparing the putative activity of upstream transcription factors. While the patient samples share activity from both ovary and testis specific TFs, there is a low activity of RFX2 and E2F6 (Figure S10B, Supporting Information). RFX2 is a as a key regulator of spermatogenesis through regulation of genes required for the haploid phase during spermiogenesis (Kistler et al., 2015), while E2F6 is critically involved in cell cycle progression (Giangrande et al., 2004). In combination with reduced MYC pathway activity, this might hint at reduced cell proliferation, which is indeed the case based on Reactome cell cycle pathways (Figure S10C, Supporting Information).

## Discussion

Because all known genetic causes of 46,XY GD were excluded by WGS, the Xp21.2 CNVs remain as the only likely cause identified in our three patients. The overlap of the *NR0B1* upstream duplicated region in P1 & P2 spans 287kb (Figure S11, Supporting Informaton) and is also contained in the triplication of P3. The 287kb region includes the genes GK and *CXorf21* together with their respective promoter and predicted enhancer regions as well as the 3′-end of TAB3 (Figure S12, Supporting Information). To our knowledge, no similar case with a Xp21.2 duplication excluding *NR0B1* or Xp21.2 triplication including *NR0B1* has been described in patients with 46,XY GD so far.

In recent years the role of three-dimensional chromosome structures and their organization in TADs have moved into the focus of genome research (Dixon et al., 2012). TADs are non-randomly organized up to megabase-sized local chromatin interaction domains that are thought to guide regulatory elements to their cognate promoters and provide insulation for non-cognate genes. These domains can be stable across different cell lines and conserved across species (Dixon et al., 2012).

TAD formation generally requires two corresponding CTCF motifs - one at either end of the loop forming domain. It has been shown that these motifs are predominantly oriented in a convergent manner (de Wit et al., 2015; Guo et al., 2015; Vietri Rudan et al., 2015) and that elimination or change of CTCF orientation can lead to removal or shift of TAD boundaries (Lupianez, Spielmann, & Mundlos, 2016; Sanborn et al., 2015). The two duplications reported in this study and the deletion reported by Smyk et al. (2007) have a 35kb overlapping region harboring a CTCF motif in proximity to the identified *NR0B1* TAD border (Figure S8 and Figure S9, Supporting Information). This CTCF is orientated towards *NR0B1*, which fits well into a concept of it being the centromeric end point of the *NR0B1* TAD (Figure S13, Supporting Information).

Hi-C data suggest a boundary region between the *NR0B1* TAD and the adjacent centromeric TAD (Figure S8, Supporting Information) (Y. Wang et al., 2018). In most cell lines this boundary region includes Exon 1 and 2 of *CXorf21* and *GK* together with their respective promoters and predicted enhancers (Figure S10, Supporting Information). Thus, *NR0B1* is shielded from these non-cognate enhancers by just a single CTCF. The discussed SVs might enable enhancers to bypass the insulating effect of this CTCF binding motif (Figure S13, Supporting Information) and allow ectopic interactions with *NR0B1*, a process known as enhancer adoption (Lettice et al., 2011) through positioning in a common loop forming domain (Figure 4). This could lead to up regulation or aberrant expression of *NR0B1*, which in consequence results in decreased SF1-mediated *SOX9* expression and impaired testicular development (Figure S2, Supporting Information). This is in accordance with the fact that all three patient’s SVs described, the deletion reported by Smyk et al. (2007), as well as previously published duplications containing *NR0B1* and *CXorf21* had maternal origin (Barbaro et al., 2012; Barbaro et al., 2007; Smyk et al., 2007).

**Figure 4:**
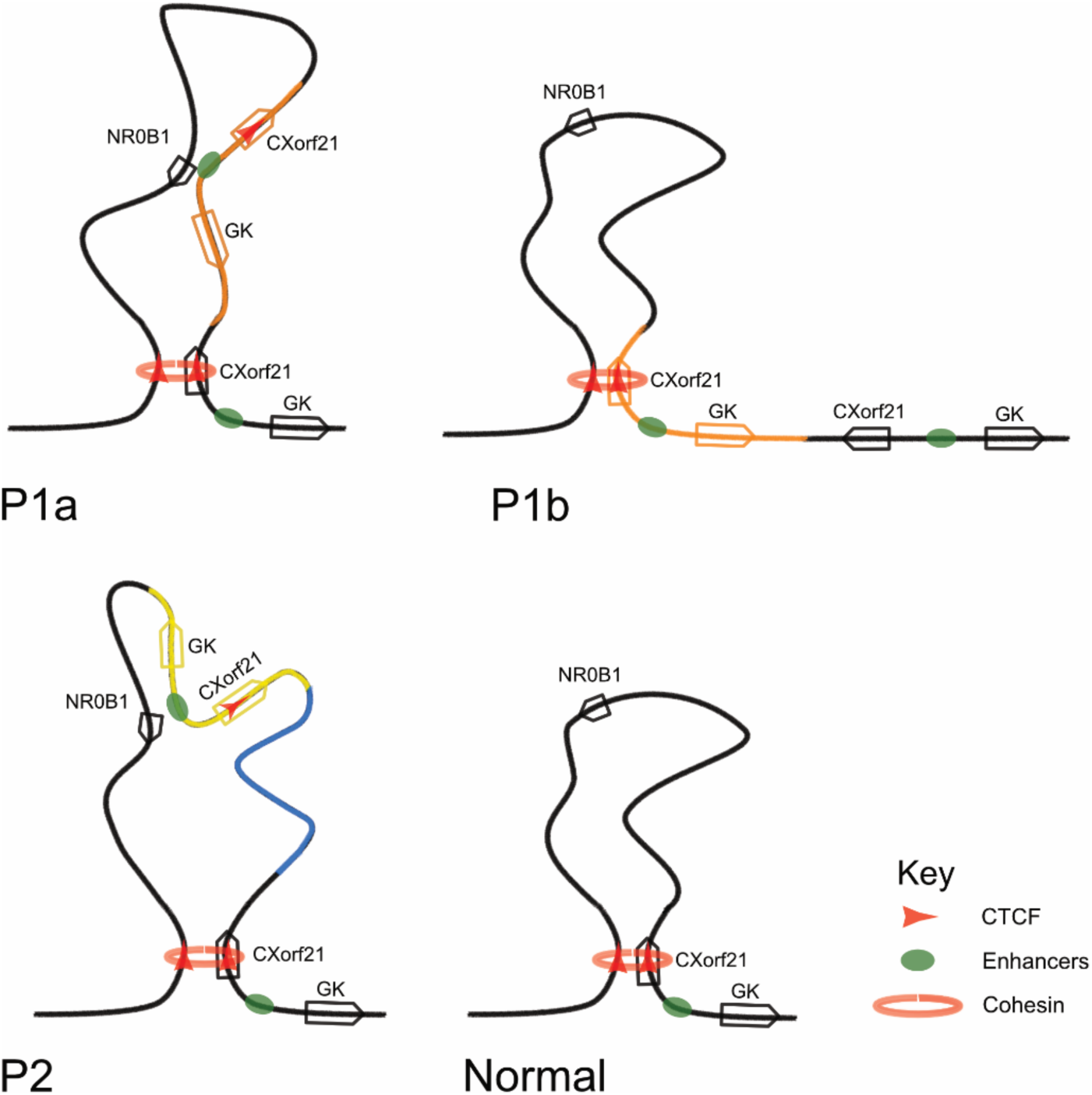
Schematic representation of topologically associating domain (TAD) in patients with *NR0B1* upstream duplications. The *NR0B1* TAD is represented as curled loop. Regions inserted through duplication are illustrated by coloured string segments. Genes are depicted by arrows showing their respective direction of transcription and are equally coloured, if part of a duplication. When a TAD comprises both the *NR0B1* and the *GK* & *CXorf21* genes with their in-between enhancers (green oval) the *NR0B1* promoter can encounter the non-cognate enhancer normally shielded away from the gene by position outside the TAD. P1a and b depict alternate TAD conformations depending on location of commenced loop extrusion.

The tandem duplication in P1 inserts an additional left oriented CTCF motif (Figure S13, Supporting Information). However, according to the cohesin-dependent loop extrusion model for the formation of TADs, only one of these two CTCF motifs can interact with the oppositely oriented CTCF that defines the telomeric *NR0B1* TAD boundary (Fudenberg et al., 2016; Sanborn et al., 2015). Loop extrusion is believed to be a dynamic process (Rao et al., 2017), whereby structural maintenance of chromosomes (SMC) complex’s (cohesin) translocate along the chromosomes in an ATP dependent process producing expanding loops of chromatin. This process brings together increasingly distant genomic loci and continues until the cohesin complex either dissociates or meets a barrier in the form of a convergent CTCF. If one side of the complex encounters a barrier it is assumed that chromatin extrusion continues on the other end until equally a convergent CTCF is met (Fudenberg et al., 2016; Goloborodko, Marko, & Mirny, 2016). Though this is a dynamic process of cohesin loading and dissociation there is an increased time of residence at TAD-boundaries (Szabo, Bantignies, & Cavalli, 2019).

The arrangement in P1 gives rise to two putative TAD conformations with either the proximal or distal *NR0B1* oriented CTCF constituting the centromeric *NR0B1* TAD boundary (Figure 4 and Figure S13, Supporting Information) with increased time of cohesin residence at both.

In situations where the *NR0B1* TAD is formed with the distal left oriented CTCF, it encompasses the whole duplicated segment which could allow the non-cognate enhancers to interact with *NR0B1* and may result in enhancer adoption (Figure 4, P1a). The identified inversion in P2 brings *GK* and *CXorf21*, along with their corresponding putative enhancers, into the *NR0B1* TAD without its inverted CTCF interfering with the existing borders (Figure 4 and Figure S13, Supporting Information). The previously outlying non-cognate enhancers are putatively inside the *NR0B1* TAD and may interact with the *NR0B1* promoter (Figure 4).

De Wit et al. (2015) also studied the deletion of individual CTCF motifs, leaving the partnering CTCF motif abandoned. Though bound by CTCF and cohesin, these formerly partnering CTCF motifs did not engage in new loops with other CTCFs. Following from this, the deletion reported by Smyk et al. (2007) most likely leads to a disruption of the TAD between the convergent sites rather than the formation of a new TAD towards a different CTCF. Several groups have studied the deletion of TAD boundaries and individual CTCF sites in different cells and organisms in recent years (Gomez-Marin et al., 2015; Narendra et al., 2015; Nora et al., 2012; Sanborn et al., 2015) showing that these result in ectopic interaction outside the previous TAD conformation. According to our model, TAD-conformation is confined to one arrangement with increased time of residence in P2 and in the patient reported by Smyk et al. (2007). The difference in the putative DNA target structures and that probability of produceing a TAD including both, *NR0B1* and the ectopic enhancers (Figure 4) could help explain the differences in phenotype of the patients. In contrast to P2, P3 and the patient reported by Smyk et al. (2007), the phenotype of P1 is less severe. The absence of a uterus in P1 implies the presence of Sertoli cells with intact production of anti-Müllerian hormone, while the other two patients (P2 and the one reported by Smyk et al, 2007) resemble a higher degree of accordance in phenotype, as both presented a small uterus and low estrogens with late onset of breast development.

In all three patients with complete gonadal dysgenesis presumed TAD-conformations bringing *NR0B1* and the non-cognate enhancers into a common domain. This is in contrast to P1, displaying partial gonadal dysgenesis, where the duplication allows a partial normal TAD conformation (Figure S7, Supporting Information). According to the loop extrusion model a multiplication of *NR0B1* and the insulator at *CXorf21* as well as the *CXorf21* and *GK* enhancers as present in P3, one of the duplicated *CXorf21* insulators loses its function due to the lack of an opposing interaction partner, whilst *NR0B1* and enhancers maintain their function with respect to the phenotype. Therefore, it is possible that this model describes a general process beyond the specific genetic rearrangements of the three patients presented here and the deletion reported by Smyk et al. (2007) and would similarly apply to all previously reported Xp21.2 duplications (Barbaro et al., 2008; Barbaro et al., 2012; Barbaro et al., 2007; Dong et al., 2016; Garcia-Acero et al., 2019; Ledig et al., 2010; White et al., 2011). It is of note, that P3 also shows muscular hypotonia possibly hinting at a regulatory effect of the triplication on the expression of the *DMD* gene.

The phenotype of Xp21.2 duplications has previously been associated with dosage effects of the duplicated genes (Barbaro et al., 2007; Bardoni et al., 1994). Recent advancements in the analysis and understanding of DNA conformation highlight the importance of intact TADs for unaltered enhancer-promoter interactions (Franke et al., 2016; Lupianez et al., 2016). With our newly discovered cases, it is not *NR0B1* itself as the unifying cause in all Xp21.2 copy number gains associated with 46, XY GD, but instead it is the 35kb upstream region, which is present in every reported case thus far (Figure 1, Supplemental Information). This underlines the necessity to reevaluate all previously reported Xp21.2 duplications, as the fundamental mechanism might be a different one then previously assumed.

Further, knowing the position and orientation of SVs alongside the new arrangement of enhancers and genes could become increasingly relevant for understanding genotype-phenotype relationship. This underscores the potential diagnostic power of whole genome sequencing, which allows accurate characterization of SVs through interrogation of individual reads at breakpoints as shown in this study.

As a result of the gene duplication, the transcriptomes of the two patients obtain an intermediate gene expression state between ovary and testis samples. Gene expression analysis clearly clusters adult ovarian and testicular tissues apart and separate from other organ tissues. The two patient samples cluster in between the two tissues, yet closer to testis and particularly close to patient tissues with low spermatogenesis. This interemediate state is further corroborated by the expression of sex determination genes (Figure S3, Supporting Information). Particularly *FOXL2* and *DMRT1*, which are hallmarks of ovarian and testicular tissue, are expressed at an intermediate level, which might be either a cause or the effect of deregulation of multiple sex determination regulators in a patient specific manner. This finding is confirmed by the gene set variation analysis using the hallmark gene sets. Both P1 and P2 show a greatly reduced expression of spermatogenesis-related genes. The latter might stem from the low expression of the transcription factor Regulatory Factor X2 (RFX2) (Figure S10B, Supporting Information), which is not only a major component of spermatogenetis in human and mouse, but also regulates cell cycle (Horvath, Kistler, & Kistler, 2004; Kistler et al., 2015). In line with this we see low activity of the upstream cell-cycle related E2F transcription factor 6 (E2F6) and cell cycle pathways are reduced in activity relative to both healthy ovary and testis tissue (Figure S10C, Supporting Information). Interestingly, MYC pathway activity is low compared to both the ovarian and the testicular tissue. MYC controls vertebrate development and contributes to developmental disorders (Hurlin, 2013). One might speculate whether the Xp21.2 duplications cause a hold of development of all or a subset of gonadal tissue cells, which is reflected by the findings from the transcriptome analysis, but which will require further in depth investigation and in vivo/In vitro confirmation.

In summary, our results demonstrate the relevance of genetic analysis beyond primary candidate genes and emphasize the importance of including more detailed analyses of genetic alterations affecting non-coding regulatory elements to routine diagnostics. This could be an important finding for uncovering novel genetic causes and understanding pathogenesis in DSD, and for patients with other disorders of unknown etiology. The findings further illustrate limitations of current diagnostic routines that are aCGH or MLPA based, as the obtained data are insufficient to evaluate most of the duplications of unknown significance. Much more research will be necessary to understand mechanisms of insulation and how regulatory elements find targets within TADs in order to completely apprehend the effect of SVs and also to make full use of techniques such as WGS in the context of sex development and far beyond.

## Data Availability

Data have not been depositet at a controlled Access database Server, yet

## Acknowledgments

The authors are truly grateful to the patients and families of these patients who cooperated in this study. The collaboration was based on the COST Action BM1303 DSDnet. H.B. and A.K. acknowledge computational support from the Lübeck OmicsCluster. This work is part of the doctoral thesis of J.A.M and A.P.S..

## Notes

**Grant Numbers** This work was funded by financial support from Bundesministerium für Bildung und Forschung BMBF (01DQ17004), Deutsche Forschungsgemeinschaft (DFG, German Research Foundation) under Germany’s Excellence Strategy (EXC 22167-390884018), Fundação de Amparo à Pesquisa do Estado de São Paulo (#2015/04763-4) and a scholarship from Coordenação de Aperfeiçoamento de Pessoal de Nível Superior (Programa de Doutorado-sanduíche no Exterior – PDSE - 88881.135162/2016-01).

### Competing Interest Statement

The authors have declared no competing interest.

### Funding Statement

This work was funded by financial support from Bundesministerium für Bildung und Forschung BMBF (01DQ17004), Deutsche Forschungsgemeinschaft (DFG, German Research Foundation) under Germanys Excellence Strategy (EXC 22167-390884018), Fundacao de Amparo a Pesquisa do Estado de Sao Paulo (#2015/04763-4) and a scholarship from Coordenacao de Aperfeicoamento de Pessoal de Nivel Superior (Programa de Doutorado-sanduiche no Exterior - PDSE - 88881.135162/2016-01).

